# OLFACTORY TRAINING EFFICIENCY IN POST-COVID-19 PERSISTENT OLFACTORY DISORDERS

**DOI:** 10.1101/2022.02.27.22271572

**Authors:** Clair Vandersteen, Magali Payne, Louise-Émilie Dumas, Élisa Cancian, Alexandra Plonka, Grégoire D’Andrea, David Chirio, Élisa Demonchy, Karine Risso, Florence Askenazy-Gittard, Charles Savoldelli, Nicolas Guevara, Philippe Robert, Laurent Castillo, Valeria Manera, Auriane Gros

**Author notes:** CORRESPONDING AUTHOR Dr. Clair Vandersteen, M.D, ENT surgery departement of Institut Universitaire de la Face et du Cou (IUFC), 31 Avenue de Valombrose, Centre Hospitalier Universitaire (CHU), Université Côte D’Azur (UCA), 06100 – NICE – France, Mail. **FUNDING** None declared.

## Abstract

**Background:** Persistent post-viral olfactory disorders (PPVOD) are widely reported after a COVID-19 and estimate to 30% one year after infection. Parosmias are the main qualitative dysosmia associated with olfaction recovery. No treatment is, to date, significantly efficient on PPVOD except olfactory training (OT). The main objective of this work was to evaluate OT efficiency on post-COVID-19 PPVOD.

**Methods:** Consecutive patients consulting to the ENT department with post-COVID-19 PPVOD were included after mainly clinical examination, the complete Sniffin’ Stick Test (TDI), the short version of the Questionnaire of olfactory disorders and the SF-36. Patients were trained to practice a self-olfactory training (professional manufactured olfactory training kit) twice a day for 6 months before coming back and undergo the same complete evaluation.

**Results:** Forty-three patients were included and performed 3,5 months of OT in average. There was a significant improvement in the mean TDI score increasing from 24,7 (±8,9) before the OT to 30,9 (±9,8) (p<0,001). Parosmias increased significantly from 8 (18,6%) to 27 (62.8%) (p<0,001). Based on normative data divided by sex and age, a significant increase in the number of normosmic participants was only found for the Threshold values (p<0,001). Specific and general olfaction-related quality of life improved after the OT.

**Conclusions:** OT seems to be efficient in post-COVID-19 PPVOD, probably on the peripheral regenerative part of the olfactory recovery. Future therapeutic strategies may focus on the central aspects of the post-COVID-19 PPVOD.

## INTRODUCTION

Long persistent olfactory complaint is widely reported after an acute, mild, or moderate, COVID-19 infection. Indeed, a complete but subjective olfaction recovery is only reported in 40 to 63%^(1–3)^ and 70%^(4)^ of patients, respectively 6 and 12 months after COVID-19. Interestingly, olfactory psychophysical tests results are better than subjective smell assessments, showing 73% to 95% normosmic patients after 6 months^(5,6)^. Some authors suggested that these remaining olfactory complaining patients, with no recovery 18 months after acute phase^(7)^, could be permanently impaired. Parosmias are the main qualitative dysosmias associated with COVID-19 olfactory recovery and occurs in 18% to 49%^(3,7,8)^ of patients 2,5 months after the acute phase of infection. Parosmias affects 20% of normosmic patients^(3)^ and contributes to the discrepancy between subjective impairment and olfactory psychophysical tests.

Long lasting olfactory loss leads to a quality of life(QoL)^(8)^ impairment, bad diet habits^(9)^, changes in social and personal relations^(10)^, psychiatric disorders (such as depression^(11)^), anxiety or anorexia^(12)^ and its nutritional consequences^(13)^, cognitive impairment^(10)^, or increase of hazardous events incidence^(14)^. Thus they have to be managed. Many treatments^(15)^ have been tried to get an olfaction recovery without significant results, including vitamins, minerals, corticosteroids, as well in COVID-19 as reported in a recent Cochrane living review^(16)^. Meanwhile, olfactory training (OT), as described by Pr. Thomas Hummel^(17)^, remains the best treatment for persistent post-viral olfactory disorder (PPVOD)^(18)^. Indeed, in PPVOD, OT systematically improve the Minimal Clinically Important Difference (MCID) of the complete Sniffin’ Stick Test score (TDI; +6^(17)^) in 30 to 68% of cases (OR=2.77)^(18)^. However, the effectiveness of OT on COVID-19 patients with PPVOD is still unknown. The main objective of this work was to evaluate OT on PPVOD after a COVID-19.

## MATERIAL AND METHODS

### Population

The study was approved by the institutional review board of the Nice University Hospital (CNIL number: 412) and registered with a ClinicalTrials.gov number (ID: NCT04799977). Since March 2020, we prospectively enrolled patients at ENT division of Nice University Hospital until December 2020. All were contaminated by COVID-19 with persistent olfactory disorders lasting more than 6 weeks (3 to 15 months). Patients where mainly self-referred or referred by general practitioners or colleagues. Patients had either a RT-PCR-proven SARS-CoV-2 diagnosis or a CT-proven SARS-CoV-2 diagnosis secondarily confirmed by serology. We retroprospectively extracted patients’ demographic data and clinical features including subjective taste impairment, subjective olfactive impairment (qualitative and quantitative dysosmia), a visual analogue scale (VAS) for the subjective assessment of olfactory recovery (ranging from 0% to 100%), weight (measured at home in the previous week on a personal scale), a nasofibroscopy (assessing nasal cavity patency and differential diagnosis), an evaluation of olfactory loss using Sniffin’ Sticks Test^®^ (SST)^(19)^, the completion of the French short version of *Questionnaire of Olfactory Disorders* (Short-QOD-NS)^(20)^ and the completion of the French SF-36^(21)^. Patients were trained to a daily use of the OT protocol (detailed below) for 6 months. A second consultation (6 months ± 15days) after OT allowed to assess a second time all the same elements except nasofibroscopy. We finally retrieved OT compliance through a score calculated according to this formula: (((number of OT weeks done/24) – (number of OT sessions missed per week / 14))/ number of OT weeks done) * 100.

### Objective olfactory dysfunction

Olfactory function was assessed using Sniffin’ Sticks test, a validated psychophysical test that included a phenyl-ethyl alcohol (PEA) odor Threshold detection (T), an odor Discrimination (D) and an odor Identification (I) test. The detailed procedure has already been detailed previously^(22)^. Results from the three tests were summed up to a composite score, the “TDI”. As described by the last update of TDI normative values^(19)^, normosmia, hyposmia and anosmia was respectively defined by a TDI≥30.75, 30.5≥TDI≥16.25, and TDI≤16. Concerning isolated T, D and I values, a normal or reduced olfactory function, related to gender and age, was respectively defined as ≥10th percentile and <10th percentile subdimension score based on^(19)^.

### Olfactory training

OT was here based on Hummel protocol^(17)^, whose regenerative properties on olfactory neurons, olfactory cortex connectivity and olfactory scores are widely reported in the literature^(15,18,23)^. Therefore, the protocol was explained to patients. It involved 6-months olfactory training with daily odors exposure, twice a day (2 sessions), with 2 different random odors of the kit, the morning and the evening (four different odors per day). We decided to run the olfactive training for 6 months as it is described^(24)^ as more effective than 12 weeks firstly described protocol^(17)^, especially since odors are renewed every 3 months^(15)^. Patients were well-informed, once all odors used at least one time, to try to recognize by blindly sniffing them. To improve compliance and ludic aspects of the OT, we used other odors than the 4 common ones (Phenylethyl alcohol [Rose], Eucalyptol [Eucalyptus], Citronellal [Citronella], Eugenol [Clove])^(17)^. We used an olfaction training kit, produced specially for this purpose by local industry, including 11 small pots of scented wax (10g), impregnated with 15% of dill, thyme, cinnamon, cloves, coriander leaf, vinegar, cumin, lavender, coffee, vanilla, or mint. We used different type of odors because there is no significant difference in olfaction improvement using simple or complex odors, or combination of both^(25)^.

### Olfactory quality of life

The olfactory QoL was assessed using the French validated Short-QOD-NS^(20)^ self-questionnaire (2 min) which is based on negative statements from the Questionnaire of Olfactory Disorders (QOD), but shorter, allowing an increase of the response rate and reducing the patient’s mental load when completing the questionnaire. These negative statements of QOD has been shown to be more correlated with the results of psychophysical olfactory tests (SST)^(26)^. The Short-QOD-NS^(27)^ evaluate the 7 most relevant questions related to social aspects (n = 3), eating (n = 2), anxiety (n = 1) and annoyance (n = 1) following an olfactory loss. The score ranges from 0 to 21 (21 meaning there is no disorder).

The 36-item form health survey (SF-36) is one of the most widely used generic questionnaires, validated in French by Perneger et al.^(28)^, used here to evaluate general QoL. The SF-36 questionnaire consists on 36 self-administered questions (5min) divided in eight domains, covering both physical and mental health. The physical component summary covers 4 subdomains as initially described: *physical functioning, role physical functioning, bodily pain*, and *general health*. The mental component summary covers 4 other domains: *vitality, emotional functioning, social functioning*, and *mental health*. The sum of the score is calculated for each domain and scaled on 100. High scores indicate good QoL while low scores indicate low QoL.

### Statistical Analysis

To explore the evolution of quantitative variables (e.g., TDI scores) before and after the OT, we employed Wilcoxon signed-rank tests, as most of the data did not follow a normal distribution (as confirmed by Shapiro-Wilks tests). To compare the evolution of binary variables (e.g, presence of parosmia, phantosmia), we employed McNemar test. To compare quantitative variables (such as TDI scores) between different groups of participants (e.g., participants with vs without parosmia) we employed Mann-Whitney tests. Non-parametric correlations (Spearman rho) were employed to investigate correlations between treatment compliance and improvement in TDI scores. All results were considered statistically significant for a bilateral alpha level of 0,05.

## RESULTS

### Demographic and clinical features

Forty-three patients were included in the study. The demographic and clinical initial features are reported in Table 1.

**Table 1.**
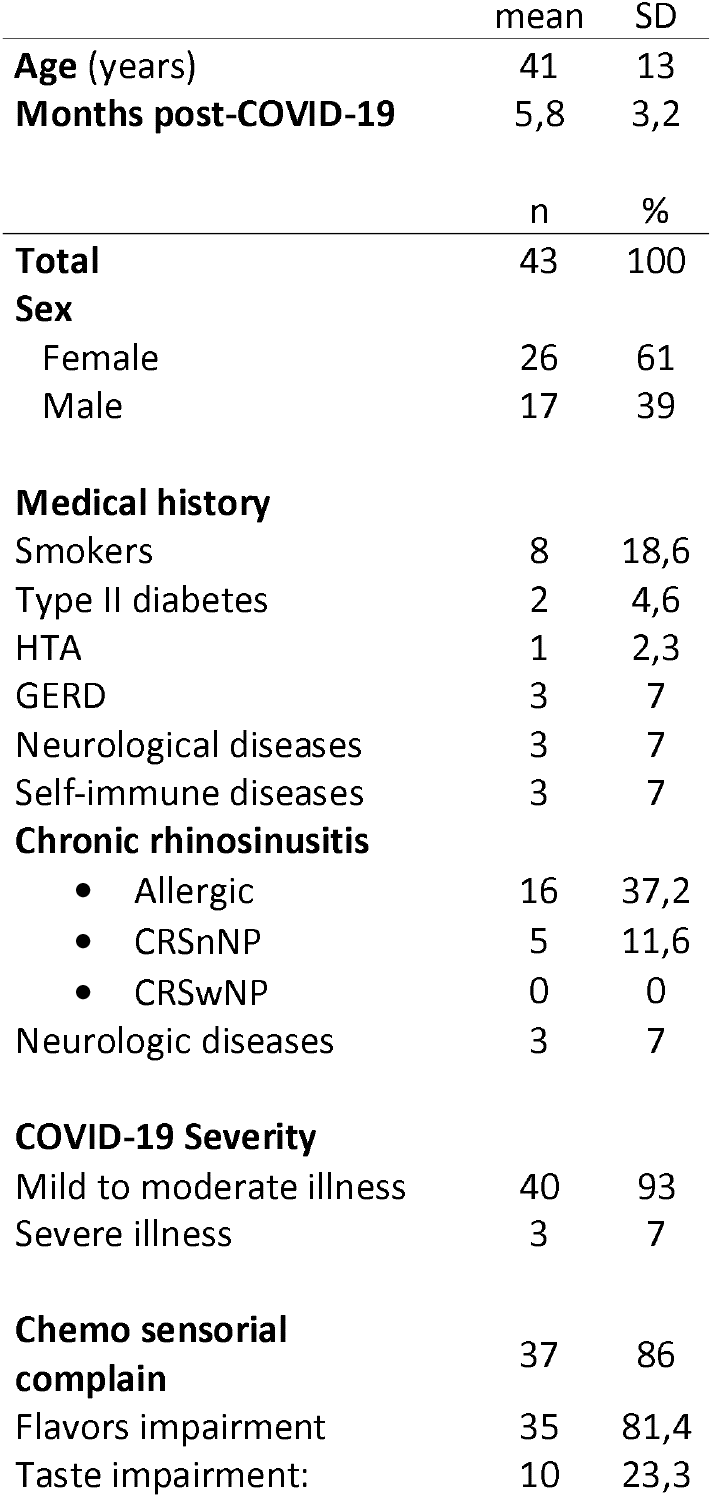
Demographics and initial clinical patients features. HTA=hypertension; GERD=gastroesophageal reflux disease; CRASnNP=chronic rhinosinusitis without nasal polyps; CRASwNP=chronic rhinosinusitis with nasal polyps

Patients were seen 5,8±3,2 months and 11±3,7 months after COVID-19 infection respectively at the first and the second (after OT) visit. Twenty-eight patients received a COVID-19 related treatment (of which 6 and 2 took respectively oral and nasal corticosteroids from 1 to 3 weeks). Among people who had a medical history of self-immune diseases, 2 had Crohn’s disease and 1 had ankylosing spondylitis. Some had a medical history of neurological diseases, 2 had epilepsy under specific medications, 1 had stroke during childness with no sequelae. Nasofibroscopies found no obstructive pathologies in the olfactory cleft.

On average, patients lost weight between the 2 visits before and after OT, going from 69,8±13,3kg to 66,7±20,1kg, but the weight reduction was not statistically significant (Z=−0,88, p=378). VAS Subjective olfactory recovery significantly increased from 34,6±26,5% to 57,9±31,1% (Z=−4,71, p<0,001). A slight, but not significant, decrease from 37 to 32 patients (74,4%) of chemosensorial complaints was reported after OT (p=0,125), with 30 (69,8%) and 5 (11,6%) patients who still suffered from flavors and/or taste loss after the OT.

### Olfactory training results

#### TDI

These was a significant improvement in the mean TDI score (Z=−4,71, p<0,001), which increased from 24,7 (±8,9) before the OT to 30,9 (±9,8) after the OT. A significant change in the number of participants categorized as anosmic, hyposmic and normosmic before and after the training was found (Chi^2^= 25,7, p<0,001). Specifically, the number of anosmic participants decreased from 10 (23,3%) to 5 (11,6%); the number of hyposmic decreased from 22 (51,2%) to 11 (25,6%); and the number of normosmic participants increased from 11 (25,6%) to 27 (62,8%). These results are graphically reported in Figure 1.

**Figure 1.**
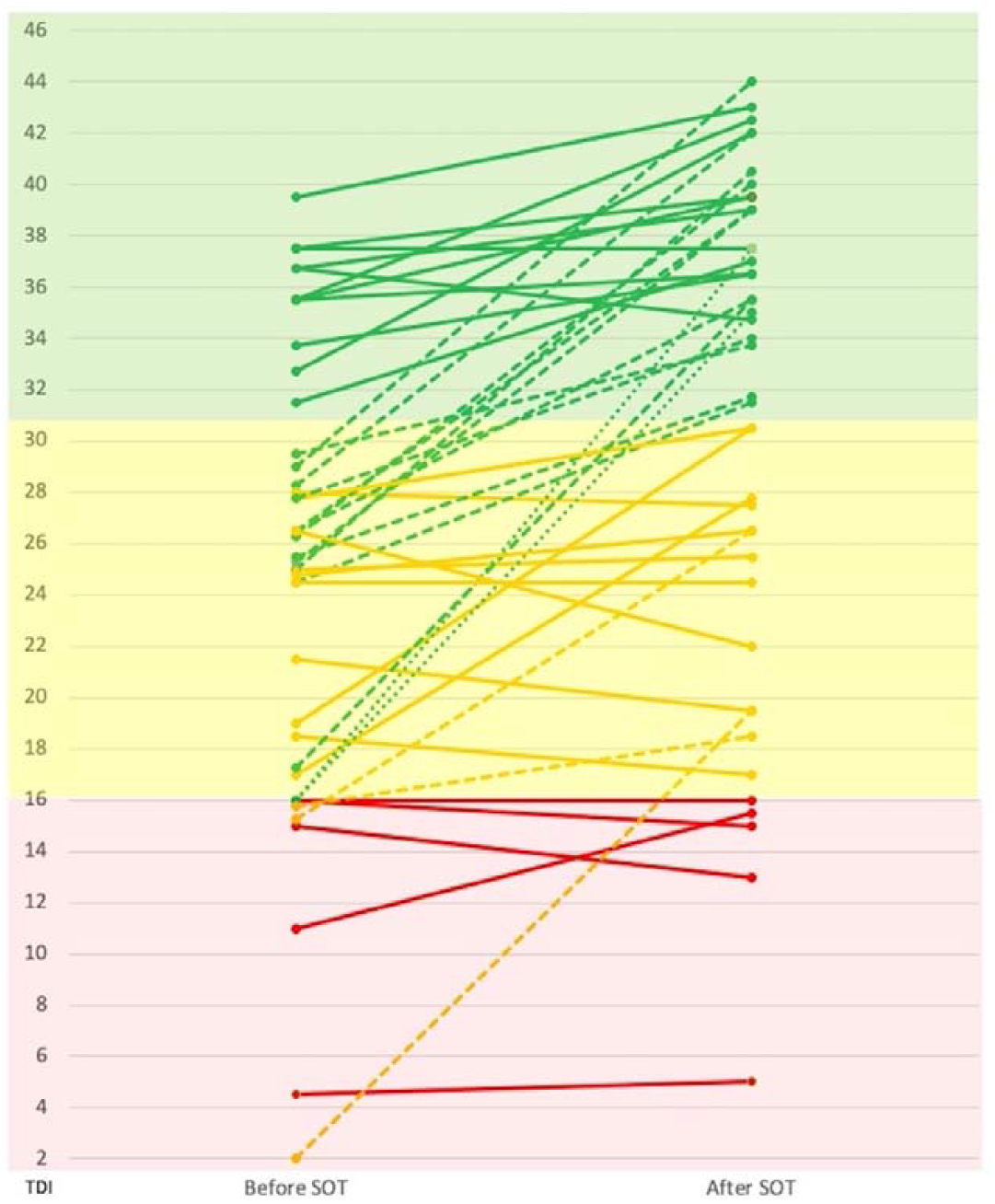
Evolution of individual TDI scores before and after OT. Colored parts cover anosmic (red), hyposmic (yellow) or normosmic (green) patients according to TDI normative values^(19)^. Oblique lines represent a patient anosmic (red), hyposmic (yellow) or normosmic (green) subject evolution according to post OT olfactory evaluation. Solid, dashed or pointed lines represent patients who respectively did not changed category, changed to the upper category, or changed from anosmic to normosmic category.

The T and I scores significantly improved after the OT (from 4,9±3,9 to 8,7±5,2, Z=−4,67, p<0,001; and from 9,4±4,1 to 11,0±3,4, Z=−3,60, p<0,001, respectively). No significant evolution of the D score was observed (from 10,4±3,0 to 11,2±3,3, Z=−1,60, p=0,110). The improvement in T was significantly bigger than the improvement in D (Z=−4,1, p<0,001) and I (Z= −2,7, p=0,007). T Improvement was significantly correlated with subjective recovery evaluation (VAS, p=0,039). Concerning the evolution of the number of participants that reached the norms for T, D, and I (based on normative datas divided by sex and age), a significant increase in normalized value was only found for the T (McNemar test, p<0,001) but not for the D (McNemar test, p=0,774) or the I (McNemar test, p=0,388). Based on age and sex normatives values, the number of participants who had normal T, D and I before and after the OT are presented in Figure 2. No clinical, medical history, treatment or compliance predictive value was significantly correlated to better psychophysical tests results.

**Figure 2.**
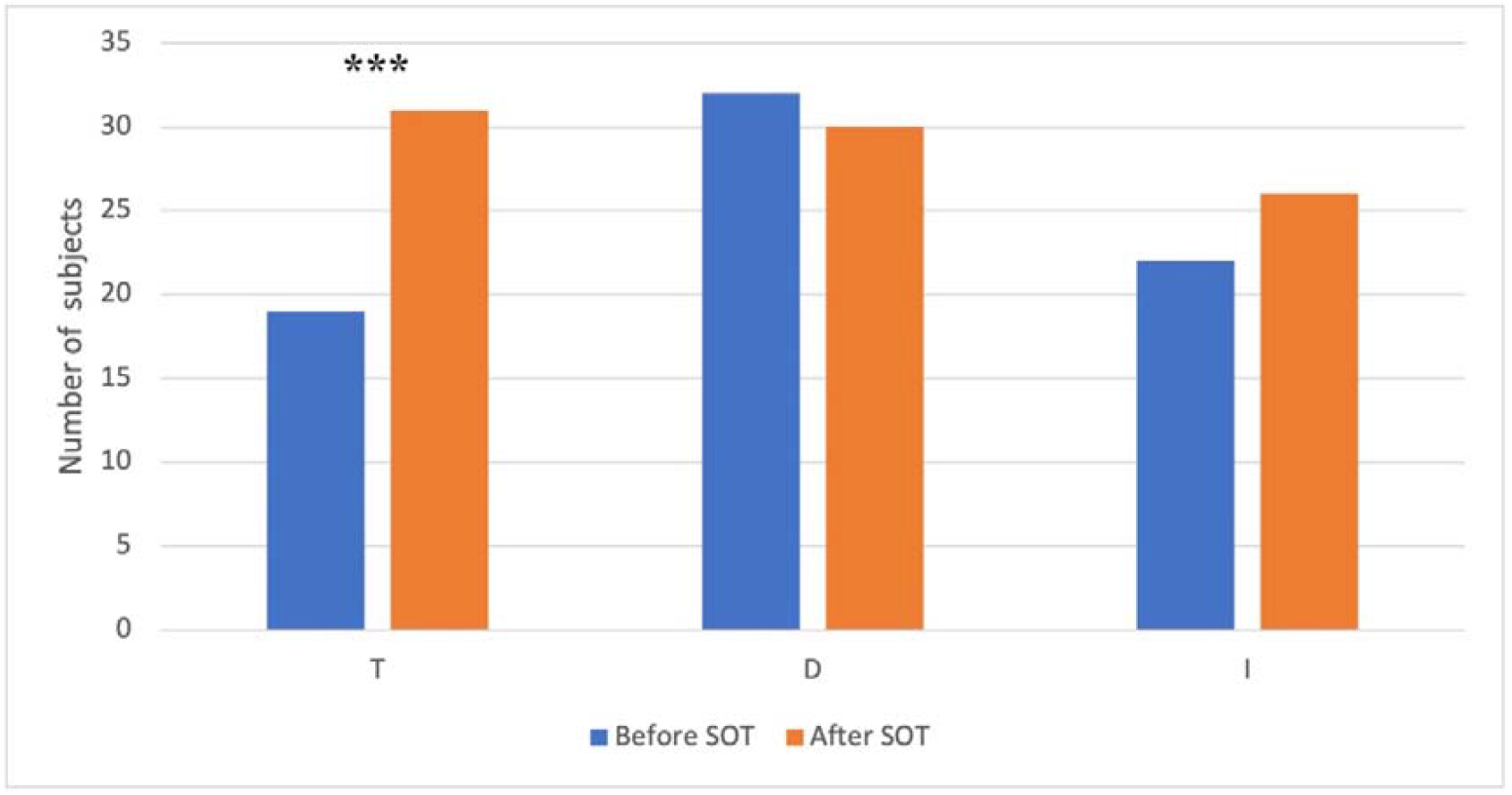
T, D & I normalization based on age and sex normative values. Number of patients who normalized T, D, and I values, before and after the OT, based on age and sex normative values.

#### Compliance

Patients performed 14,5±9 weeks (∼3,5months) of OT on average, missing 0,8±1 entire day per week, and 3±3 sessions per week. The average compliance ratio was 53±33%, ranging from 4% to 110% (for people that did more than 24 weeks). There was no significant correlation between OT compliance and TDI evolution or subjective olfaction evaluation recovery – VAS, or improvements in quality of life (all ps> 0,101).

#### Qualitative dysosmia

The number of participants reporting the presence of parosmia increased significantly from 8 (18,6%) to 27 (62.8%) after the OT (McNemar test, p<0,001), with only 1 participant (2%) that fully recovered after the OT, and 20 participants (46,5%) that developed parosmia after the OT. At the end of the OT, participants presenting parosmia showed lower identification scores (U= 122, p=0,018). Patients with parosmia were significantly less likely to lose weight (Z=−2,4; p=0,013). No significant difference in the number of participants reporting phantosmia was found (9 participants – 20,9% - before the OT, and 12 - 27.9% - after the OT, p=0,581; 8 subjects – 18,6% - developed phantosmia after the OT, and 5 – 11,6% - recovered after the OT). No predictive factor was significantly associated with qualitative dysosmia evolution.

#### Quality of life

The results of the SF36 and the Short-QOD-NS questionnaires are reported in Table 2. Concerning the SF36, significant improvements after the OT were obtained in the subdomains assessing physical functioning (p=0,009), social functioning (p=0,013=, emotional role (p=0,049), vitality (p=0,023), and general health perception (p=0,045). For the Short-QOD-NS, significant improvements after the OT were observed for the total score (p<0,001) and all the subdomains, namely the social (p=0.001), the food (p=0,036), the anxiety (p=0,020), and the annoyance (p=0,020) subdomains. Short-QOD-NS improvement was significantly correlated to a TDI improvement after OT (p=0,008).

**Table 2.** Olfactory specific (Short-QOD-NS) and general (SF-36) quality of life comparative results before and after OT. p represents the p-value at the Wilcoxon signed-rank test. *p<0,05; **p<0,01; ***p<0,001

| Quality of life Score | BEFORE OT |  | AFTER OT |  | *p* |
| --- | --- | --- | --- | --- | --- |
|  | Mean | ET | Mean | ET |  |
| **Short-QOD-NS** |  |  |  |  |  |
| *Total score* | 10,44 | 5,97 | 13,65 | 6,49 | <0,001*** |
| *Social subdomain* | 4,58 | 2,7 | 5,88 | 2,86 | 0,001** |
| *Food subdomain* | 2,98 | 2,22 | 3,72 | 2,42 | 0,036* |
| *Anxiety subdomain* | 1,91 | 1,02 | 2,44 | 1,12 | 0,020* |
| *Annoyance subdomain* | 0,98 | 1,03 | 1,60 | 1,20 | 0,020* |
| **SF36** |  |  |  |  |  |
| *Physical functioning* | 79,53 | 25,42 | 85,81 | 23,20 | 0,009** |
| *Social functioning* | 66,28 | 28,68 | 74,13 | 26,78 | 0,013* |
| *Physical role* | 66,28 | 40,42 | 71,51 | 38,80 | 0,407 |
| *Emotional role* | 55,81 | 41,61 | 64,34 | 42,04 | 0,049* |
| *General mental health* | 62,05 | 21,60 | 61,11 | 21,33 | 0,844 |
| *Vitality* | 44,54 | 24,07 | 51,86 | 21,63 | 0,023* |
| *Bodily pain* | 66,61 | 31,64 | 69,63 | 33,51 | 0,337 |
| *General health perception* | 63,63 | 26,09 | 69,56 | 23,22 | 0,045* |

## DISCUSSION

Persistent post-COVID olfactory loss is becoming a social issue as millions of people worldwide are affected. OT is, for the moment, the only therapeutic hope for post-COVID-19 olfactory impaired patients who are still complaining of it many months after contamination, despite spontaneous olfactory recovery occurring in 40^(2)^ to 70%^(4)^ of cases from 6 to 12 months. For now, only a few studies reported the OT efficiency in post-COVID-19 PPVOD.

This study reports a significant olfactory recovery after ∼3,5 months of OT in PPVOD related to COVID-19. The SST MCID increased by more than 6 points^(17)^ in average suggesting, in this situation, the OT efficiency as already knew for non-COVID-19 related PPVOD^(15,18,24,25)^. Interestingly, we observed more than a doubling normosmic patients’ ratio after OT, going from 11 (25,6%) to 27 (62,6%).

Specific to COVID-19, OT results were only reported with complete SST results in Le bon et al.^(29)^ study that compared 10 weeks of OT with (n=9) or without (n=18) a 10 days oral corticosteroids course. In this study, there was no significant olfactory recovery in OT alone group but report 2 nonhomogeneous groups and a poor compliance for 31% of patients. Olfactory subdimensions (T, D and I) specific recoveries were not reported by authors^(29)^. In COVID-19 PPVOD, OT alone was reported as significantly improving olfaction recovery only in other steroids efficiency evaluations studies but never again with a complete SST evaluation^(30,31)^. However, it is recommended^(15)^ to integrate T, D and I studies in olfactory evaluation. Indeed, OT effect on T, D and I in case of PPVOD is still unclear. Hummel firstly described a clear T increasing effect^(17)^ of OT. So, according to our results, Oleszkiewicz et al.^(25)^ reported a significant increasing effect on T and I in OT efficiency on post-infectious (n=57) and idiopathic (n=51) olfactory long lasting dysfunctions. T-recovery could be explained by a peripheral regenerative^(32)^ effect of OT with a regrowth of olfactory neurons^(33)^ ; and I-recovery (with D-recovery) by a more central processing allowing an olfaction dedicated area connectivity reorganization^(23,25)^. More recently, Sorokowska et al.^(24)^ reported in a meta-analysis (n>879) a large and significant post-OT increase both on D and I. However, they^(24)^ also mentioned a small to moderate effect on T, which is in contradiction with our results despite that we used the same PEA threshold in both visits. According to these studies^(17,24,25)^, in a PPVOD situation, we report an expected significant T-recovery compared to an insufficient I and D-recovery, the two last being usually correlated to higher olfactory functions.

As potential neurological outcomes of COVID-19 PPVOD are getting more and more discussed in literature^(34,35)^, it could be an explanation for this lack of significant I-normalization and D-improvement. Moreover, the D is the only subdimension which did not significantly improved while it’s well documented that the olfactory-hippocampal network is actively involved during a discrimination learning, and so OT^(36)^. Discrimination and identification tasks are closely related to cognition and specifically to executive functions, semantic task and episodic memory^(37)^. These cognitive functions might be affected by hypometabolism and dysfunctioning of many parts of secondary olfactory cortex areas, or areas connected to them, reported in 18FDG PET study^(38)^ on COVID-19 PPVOD such as : bilateral orbito-frontal cortex, cingulate gyrus, thalamus, hippocampic or parahippocampic gyri. Theses cognitives and semantic isolated impaired areas could be part of a more global connectivity structure impairment suggested in a tractography study which is the inferior longitudinal fasciculus^(35)^. Moreover, MRI morphological (and functional) modifications of many of these cortical areas, especially gray matter volume of cingulate gyrus and hippocampus^(39)^, are reported to be correlated to COVID-19 persistent (≥3 months) smell loss and could be the mark of a long lasting damage of dedicated olfactory areas. Currently, there is still some doubt regarding the etiology of central abnormalities observed as they are not proven to be the cause or the consequences of persistent olfactory loss, but the link between D and I impairment and impaired olfactory brain areas is becoming more and more obvious. These central involvement in post COVID-19 persistent olfactory loss may suggest a therapeutic approach consisting in the use of cognition and semantic training that could be mediated by speech therapist. It has to be evaluated in the future.

In our study, parosmias were multiplied by 3 after OT. Parosmia physiology is complex and poorly understood. It seems to be an olfactory epithelium regeneration side effect spontaneously emerging in 18%^(8)^ to 43%^(3,7)^ of COVID-19 PPVOD patients. Peripheral origin is supported by an abnormal neuronal regrowth, including bad proximity neurons contacts, in a hypotrophic olfactory bulbs environment^(40)^. Parosmias annoyance is not systematic ally correlated with olfactory function justifying it occurs sometimes after a total olfactory recovery in 2[here] to 20%^(3)^. Central origin is supported by gray matter alterations^(41)^ and olfactory cortex hypometabolism^(42)^. Thus, the increasing parosmic patient’s ratio could be linked to peripheral regeneration induced by OT, as suggested by the significant T increase. In contrast, the persistence of this symptom could be correlated to a lack of central processing suggested by the lack of I-recovery. Widely, the fact that D and I did not normalize could support a central involvement explanation for persistent post-COVID-19 olfactory loss.

Moreover, we found that parosmic patients did not lose weight, unlike non parosmic patients. Olfaction disorders is a well-known state correlated with abnormal human control of food intake portion size and decreased reward system signals and so satiety^(43)^. This study supports this effect as olfactory recovery seems to be associated with a slight but not significant loose of weight acquired during the anosmic phase. However, when parosmias occurred, this could prevent this weight loose and increase a potential metabolic risk factor associated to salt and sugar intake increased in near 30% of COVID-19 patients, especially in case of anosmia^(44)^.

Smell loss cause a well-known significant QoL worsening^(15,45)^.The benefit of such a OT in COVID 19 PPVOD in still unknown but we previously reported an alteration of Short-QOD-NS^(8)^. In this study, we reported that OT not only induced significant improvement of Short-QOD-NS, but also general QoL through SF-36 results. All Short-QOD-NS sub-scores were significantly improved but mainly one related to social relationships. After a long olfactory deprivation time, patients get used to it and develop strategies to cope with parosmias such as avoiding food tasting or not smoke smelling. These behaviors generate anxiety, and patients suffer from social network reduction. Moreover, loneliness contributes to the 30% increase of depression and suicide in this specific population^(46)^. The *emotional role* and *vitality* SF-36 sub-domain improvements (table 2) are consistent with the fact that olfaction is more than just a food sense, but also a channel for social, sexual, and emotional communication. Healing from an olfactory loss seems to improve mental general state of patients. Smeets et al.^(45)^ previously reported that all SF-36 subdimensions which improved in our study, except general mental health, were significantly impaired in case of severe dysosmia underlining the specific effect of OT on dysosmia related general QoL.

This study had however several limitations. We did not use a control group as it’s often the case in OT study because it’s ethically difficult not to treat a patient, and technically impossible or quite difficult to use placebo odors. Concerning TDI results, we can wonder if spontaneous recovery could have produced the same results. It’s hard to answer to that question formally as, with similar complete (T,D,I) SST evaluations, there are only a few studies reporting spontaneous post-COVID-19 olfactory recovery^(47,48)^, and even less reporting it after OT^(29)^. Iannuzzi et al.^(49)^ found a significant T progression evaluated after 2 months of spontaneous olfactory recovery, which may correspond to early olfactory neurons and sustentacular regeneration occurring around 2 to 4 weeks in such inflammatory environment^(50)^. Compared to D and I, we previously reported^(8)^ that T was the most decreased olfaction subdimension measured in a cohort of patients with near 6 months post-COVID-19 PPVOD, confirming what many authors found in similar patients after 4^(48)^ or 6^(47)^ months of spontaneous recovery with a tiny or non-significant increasing of T. There is, to date, no potential explanation that could validate a spontaneous T increasing after 6 months of persistent post-COVID-19 olfactory loss. Spontaneous recovery occurred mainly in the first month and does not change afterward. Another limit is the absence of psychophysical taste evaluation even if, after 6 months, almost all patients no longer complained about it. Last main limit is the small sample size which can reduced the study strength. Therefore, our SST subdimensions OT results singularity must be confirmed with larger cohorts of patients.

## CONCLUSION

In COVID-19 PPVOD, OT seems to significantly improve olfaction recovery, more than doubling normosmic patient’ ratio. Olfaction subdimensions and qualitative dysosmias studies underline the main regenerative peripheral effect of OT as olfaction threshold significantly improved. Despite of olfaction, specific, and general patients’ quality of life improvement, central involvement of persistent olfactory loss is becoming more and more significant and must be focused on future studies to improve olfactory quantitative and especially qualitative recovery.

## Data Availability

All data produced in the present study are available upon reasonable request to the authors

## ACKNOWLEDGEMENTS

Authors want to thank PAYAN BERTRAND perfumery society (28 Av. Jean XXIII, 06130 Grasse, France) who graciously manufactured olfaction training kits given to each patient in this study.

## AUTHORSHIP CONTRIBUTION

CV, MP, EC, LED, AG contributed to study design, data collection, interpretation of results, drafting and critical evaluation of the final manuscript. AP, GD, DC, ED, KR, FA, CS, NG, PR and LC contributed to study design, interpretation of results, drafting and critical evaluation of the final manuscript. VM contributed to biostatistics, interpretation of results and critical evaluation of the final manuscript.

